# Greenness, Blue Spaces and Human Health: An Updated Umbrella Review of Epidemiological Meta-analyses

**DOI:** 10.1101/2024.06.20.24309223

**Authors:** XiaoWen Wang, Bowen Feng, Juan Wang

**Author notes:** Contributed equally (XiaoWen Wang, Bowen Feng).

## Abstract

We systematically summarizes and evaluates the relationship between green and blue spaces and human health through an umbrella review of epidemiological meta-analyses up to the year 2024. Green spaces have been recognized for their ecological services, including air purification and biodiversity protection, which contribute to the enhancement of life quality and well-being. The review highlights significant advancements in research methodologies and the emergence of new evidence linking green spaces with reduced risks of various health issues, such as type 2 diabetes, obesity, cardiovascular diseases, and improved mental health.

The study follows the PRISMA guidelines and includes meta-analyses from PubMed, Embase, and Cochrane databases, focusing on new evidence and methodological improvements. Inclusion criteria encompass studies on human populations, exposure to green and blue spaces, and health outcomes such as mortality, disease risk, and physiological indicators. Data extraction and quality assessment of evidence and methods are conducted using the GRADE system and AMSTAR 2 tool.

The review finds that green space exposure is associated with reduced all-cause mortality, cardiovascular disease mortality, incidence of diabetes and metabolic syndrome, low birth weight, and mental health improvements. Blue spaces also show positive associations with health outcomes, including reduced obesity rates and improved psychological well-being. However, the evidence regarding green space exposure and specific health outcomes like cancer, asthma, and allergic rhinitis remains heterogeneous and unclear.

The review underscores the need for future research to address methodological limitations, incorporate various green space indicators, and explore the complex mechanisms of human-environment interactions. It concludes by emphasizing the importance of green and blue spaces in urban planning and public health strategies to improve residents’ health and quality of life.

## Introduction

As an essential component of the human living ecosystem, green spaces have increasingly drawn attention for their impact on human health(Taylor & Hochuli, 2017). Green spaces not only provide recreational areas but also offer various ecological services such as air purification, mitigation of the heat island effect, and biodiversity protection(Kumar, Ekka, Upreti, Shilky, & Saikia, 2023). These services play a crucial role in enhancing residents’ quality of life and physical and mental well-being. Numerous studies in recent years have confirmed the positive association between green spaces and human health, including promoting physical activity(McMorris, Villeneuve, Su, & Jerrett, 2015), reducing mortality rates(Ji et al., 2019), lowering the risk of cardiovascular diseases(Pereira et al., 2012), and improving mental health (J. Wang et al., 2024).

Although a systematic review of this field was conducted in 2021(Yang et al., 2021), significant advancements in the study of the relationship between green spaces and human health have been made over the past three years due to continuous improvements in research methodologies and the emergence of new scientific evidence. These advancements include identifying and confirming more health outcomes associated with green spaces, such as the risks of type 2 diabetes(Tsai et al., 2021; Yu et al., 2022) and obesity (Fan et al., 2022; Xiao et al., 2021). Correspondingly, new literature reviews and meta-analyses have increased not only in quantity but also in the depth and breadth of research. These studies cover various aspects, from the impact of green spaces on specific health outcomes to how green space characteristics(H. Wang & Tassinary, 2024), frequency of exposure(Xia et al., 2024), and socioeconomic factors modulate this relationship(Jamalishahni, Turrell, Foster, Davern, & Villanueva, 2023).

This study aims to systematically summarize and evaluate all meta-analyses on the relationship between green spaces and human health up to 2024. 6 through an umbrella review of evidence provided by epidemiological studies(Choi & Kang, 2023). We focus on new evidence, improvements in research methods, potential differences, and controversies to supplement and update the existing knowledge system. Additionally, we will update the assessment of another important system in the living environment: the impact of blue spaces on human health. We aim to provide the latest scientific evidence for public health decision-makers, urban planners, and environmental protection policymakers, guiding them in formulating more effective strategies to promote the protection and utilization of green spaces, thereby improving the health and quality of life of urban residents, and to anticipate future research directions.

## Methods

We conducted a systematic umbrella review of the meta-analyses following the Preferred Reporting Items for Systematic Reviews and Meta-Analyses (PRISMA) guidelines(Page et al., 2021). The protocol for this umbrella review has been registered in the International Prospective Register of Systematic Reviews (Prospero), ID: CRD42024533346.

### Inclusion criteria and searches

We systematically searched three international electronic databases: PubMed, Embase, and Cochrane. Our search strategy used terms related to green spaces (“green space,” “green spaces,” “greenspace,” “greenspaces,” “greenness,” “normalized difference vegetation index,” “soil-adjusted vegetation index,” “enhanced vegetation index,” “vegetation,” and “leaf area index”), blue spaces, gardening, forest bathing, and exposure to natural environments, as well as systematic reviews and meta-analyses (“systematic review” or “meta-analysis”). We included studies published up to May 30, 2024, and restricted our search to research articles. We also manually cross-checked the results of the title and abstract searches to remove duplicates.

Two researchers (W.X. and F.B.) independently screened the titles and abstracts to determine study inclusion. Discrepancies were resolved through discussion with a third author (W.J.). Our inclusion criteria were as follows: (1) Population—studies on human populations regardless of age, gender, race, geographic region, and health status; (2) Exposure—studies on exposure to green and blue spaces, including residential green spaces (assessed using vegetation indices, proportion of green space, proximity to green spaces, or the amount of green space in a specific area) and activities conducted in natural environments (e.g., exercising in nature, gardening); (3) Comparison—studies comparing health impacts of different levels of green space exposure; (4) Outcomes— studies investigating any health outcomes, such as mortality, disease risk, prevalence, incidence, and physiological indicators. We did not impose any design restrictions on the primary studies included in the meta-analyses. We excluded original studies, non-human studies, and conference abstracts. Articles published in languages other than English were also excluded.

### Data extraction

Two authors (W.X. and F.B.) independently extracted the data, with discrepancies resolved through discussion with a third author (W.J.). For each eligible systematic review, the following information was extracted: author, publication year, study design (observational/interventional), main findings, and characteristics of the included primary studies. These characteristics included age range, sample size, methods of assessing green/blue spaces, health outcomes, effect sizes, 95% confidence intervals, and statistical significance.

### Credibility and quality assessment of evidence and methods

We used the GRADE (Grading of Recommendations, Assessment, Development, and Evaluation) system to assess the quality of evidence for each outcome in each meta-analysis, categorizing them as “high,” “moderate,” “low,” or “very low” (Guyatt, Oxman, Schünemann, Tugwell, & Knottnerus, 2011). According to GRADE standards, all observational studies are considered low-quality evidence. The GRADE method includes eight criteria, five of which can lower confidence in the accuracy of effect estimates, resulting in downgrading: risk of bias, inconsistency of results, indirectness of evidence, imprecision, and publication bias. Additionally, three criteria can increase or enhance confidence: a large magnitude of effect with no plausible confounders, a dose-response gradient, and all plausible residual confounders would reduce the effect or suggest a spurious effect if not controlled. Two authors independently assessed each item based on the content of the articles. Heterogeneity was primarily evaluated using the I² value: we defined 0-30% as low, 30-70% as moderate, and above 70% as high heterogeneity. If high heterogeneity was observed, the evidence was downgraded.

We used AMSTAR 2 to assess the quality of individual meta-analyses. The quality of included meta-analyses was evaluated using AMSTAR 2 (A Measurement Tool to Assess Systematic Reviews – second edition) (Shea et al., 2017). Two authors independently assessed each item of the tool, and any discrepancies were discussed with a third author. The AMSTAR2 checklist contains 16 items that include questions related to (1) the use of the PECO framework as study question or inclusion criteria, (2) a priori protocol for the review, (3) the selection criteria for the study design, (4) the comprehensiveness of the literature search strategy, (5) the number of authors performing the literature selection, (6) the number of authors performing the data extraction, (7) the reporting of the characteristics of the excluded studies, (8) the reporting of the characteristics of the included studies, (9) the risk of bias assessment for the included studies, (10) the reporting of the sources of funding in the included studies, (11) the use of appropriate statistical methods for any meta-analyses reported, (12) the impact of risk of bias in the included studies on the pooled results, (13) the explanation for the risk of bias in the included studies and its impact on the results of the review, (14) the explanation for the heterogeneity in the review, (15) the investigation of publication bias, and (16) the reporting of conflicts of interest in the review. Of these, items 2, 4, 7, 9, 11, 13, and 15 were identified as critical domains and the remaining were considered non-critical domains by the authors checklists.

## Result

### 1. Systematic review retrieval

The initial search identified 4,474 records. After removing duplicates, 4,213 titles and abstracts of systematic reviews were assessed, and 4,125 articles were excluded during the title and abstract screening. A total of 88 articles were subjected to full-text review. Of these, 34 articles were further excluded for being irrelevant to the topic or focusing on other priorities, 2 articles(Jia et al., 2023; Patwary et al., 2022) were excluded due to unavailability of full text or being conference abstracts, and 6 articles (Annesi-Maesano et al., 2023; Gascon et al., 2016; A. Lambert, Vlaar, Herrington, & Brussoni, 2019; M. Liu et al., 2023; Moore et al., 2018; Ye et al., 2022)were excluded for not being quantitative analyses. Finally, 46 meta-analyses were included in the umbrella review.

### 2. Characteristics of systematic reviews included in the umbrella review

The umbrella review included 46 systematic reviews with meta-analyses. The majority of these articles were published between 2021 and 2024, with 33 articles (approximately 72%) published after 2021 (Table 1). The number of primary studies included in the meta-analyses for each health outcome ranged from 2 to 76. Most of the included primary studies were observational, followed by experimental/intervention studies. The study populations covered all age groups, from infants to the elderly, and the research mainly focused on countries and regions capable of conducting large cohort studies, such as North America, Europe, and China.

### 3. Greenspace exposure measures

In recent studies, various metrics have been used to assess green space exposure. Objective parameters include the Normalized Difference Vegetation Index (NDVI) (29/46), Leaf Area Index (LAI) (1/46), area of green patches (6/46), distance to the nearest green space (2/46), and the number of nearby parks (1/46). One quantitative review analyzing the health impacts of residential building characteristics included descriptions of land use types (1/46). Subjective parameters included self-reported exposure and visits to natural environments (8/46). We separately assessed activities conducted in natural environments, such as forest bathing (2/46) and gardening (4/46). Blue space exposure assessments included distance, the presence of blue spaces within specific buffer zones, blue space coverage, and self-reported frequency of use.

### 4. Health outcomes

We categorized the health outcomes related to green space exposure into several sections: outcomes associated with green space exposure include mortality, neurological disorders and cognitive function, cardiovascular and metabolic diseases (including CVD, diabetes, metabolic syndrome, overweight and obesity, metabolic indicators), cancer, allergic diseases (mainly affecting children and adolescents), pregnancy outcomes, and mental health. Additionally, we conducted a separate review of blue space outcomes. These health outcomes were measured using various methods, including physician diagnosis, questionnaire surveys, records from hospitals or other health-related departments, self-reported health status, and laboratory tests.

### 5. Methodological quality

Many of the included systematic reviews did not meet all seven key domains of the AMSTAR 2 checklist (Table 1). 24 out of 46 reviews (52%) developed a protocol for this review. 45 out of 46 reviews (98%) conducted a comprehensive literature search or pre-specified specific cohorts. 11 out of 46 reviews (24%) provided a list of excluded studies and demonstrated the rationale for exclusions. 45 out of 46 reviews (98%) used appropriate meta-analysis methods. 42 out of 46 reviews (95%) considered the risk of bias in primary studies when discussing the results of the systematic review or pre-specified specific cohorts. However, only 32 out of 46 reviews (70%) assessed the risk of small sample bias (publication bias) in primary studies. We used GRADE grading to assess each study’s evidence level for each health outcome (N=154). The majority of the evidence from studies ranged from “very low” to “low” quality.

### 6. Associations between greenspace exposure and health outcomes

#### 1. Mortality

Several systematic reviews and meta-analyses have shown a significant association between green space exposure and reduced all-cause mortality. For the general population, an increase of 0.1 unit in NDVI around residential areas is associated with a 4% to 7% reduction in all-cause mortality risk(Bertrand, Pascal, & Médina, 2021; Rojas-Rueda, Nieuwenhuijsen, Gascon, Perez-Leon, & Mudu, 2019). Among the elderly population, each 0.1 unit increase in NDVI is linked to a 1% reduction in all-cause mortality risk(Yuan, Huang, Lin, Zhu, & Zhu, 2021). Additionally, green space exposure may reduce disease-specific mortality rates by providing a healthier living environment. For instance, every 0.1 unit increase in NDVI is associated with a 2-3% reduction in cardiovascular disease (CVD) mortality, ischemic heart disease (IHD) mortality, and cerebrovascular disease (CBVD) mortality(Bianconi, Longo, Coa, Fiore, & Gori, 2023; Liu et al., 2022; Twohig-Bennett & Jones, 2018). Other quantitative analyses found beneficial associations between green space and mortality rates related to neurodegenerative diseases(F. Li et al., 2023) and chronic obstructive pulmonary disease(M. Tang, W. Liu, H. Li, & F. Li, 2023). For the elderly, each 0.1 unit increase in NDVI corresponds to a 23% to 33% lower risk of stroke mortality(Yuan et al., 2021). Two studies explored potential associations between green space exposure and cancer mortality, with quantitative analyses suggesting potentially beneficial associations with lung cancer and prostate cancer mortality(J. Li et al., 2023; Sakhvidi et al., 2022). These studies consistently provide evidence (GRADE: III-V) demonstrating the beneficial effects of green space exposure on overall health risks in the general population, particularly regarding cardiovascular disease mortality. However, studies on mortality risks related to other diseases currently lack quantitative data, and the evidence quality is very low, necessitating cautious interpretation of the study results.

#### 2. Neurological disorders and cognitive function

Recent studies have highlighted the association between green space exposure and neurological system diseases (NSD), which is a significant concern in public health. A meta-analysis covering 15 studies investigated the relationship between greenness exposure and NSD outcomes, including cerebrovascular diseases, stroke, and neurodegenerative diseases(F. Li et al., 2023). The analysis found a significant negative correlation between greenness exposure and the risk of NSD mortality or incidence/prevalence.

Specifically, two studies observed that green space exposure could be a protective factor against dementia among various environmental exposures in residential settings(Yimin Zhang et al., 2024; Zhao et al., 2021). However, a dose-response study separately examined the association between greenness and dementia. It found a slight negative correlation at moderate levels of greenness exposure but no association at high levels(Zagnoli et al., 2022).

While evidence remains limited, factors related to climate-related exposures, including air pollution(Dong et al., 2021), short-term extreme heat(Yuzi Zhang et al., 2023), and climate change(Bongioanni et al., 2021), may exacerbate symptoms of Alzheimer’s disease and related dementias (ADRD) and Parkinson’s disease (PD), and disproportionately affect them. Exposure to green spaces, vegetation, or parks may mitigate the impacts of these exposures. However, existing studies are limited and inconsistent, suggesting very low levels of evidence (GRADE: IV-V).

Commonly used green space metrics may not capture specific outdoor green space utilization, and less investigation into policy-related and socio-economic protective characteristics, such as economic development status and education level, remains. These overlooked features could be crucial factors influencing how green space exposure mediates neurological and cognitive function.

#### 3. Cardiovascular and metabolic diseases

The relationship between green space exposure and cardiovascular diseases and metabolic health has garnered considerable attention. Previous studies indicate that meta-analyses consistently show that green space exposure reduces mortality rates associated with ischemic heart disease and cerebrovascular diseases, though evidence regarding disease incidence is inconsistent. Research suggests that green spaces can lower the risk of cerebrovascular diseases(F. Li et al., 2023), but evidence regarding the impact on cardiovascular disease risk is limited(Twohig-Bennett & Jones, 2018).

Regarding type 2 diabetes mellitus, different studies indicate that greater exposure to green spaces is associated with reduced diabetes risk(Den Braver et al., 2018; Sharifi et al., 2024; Twohig-Bennett & Jones, 2018), potentially linked to higher community walkability(Den Braver et al., 2018). Multiple systematic reviews and meta-analyses have explored the relationship between green space and metabolic health factors, including obesity(Luo et al., 2020), body mass index (BMI), hypertension (HTN) (Bu et al., 2024), blood glucose (BG), and lipid profiles(Sharifi et al., 2024). Studies indicate that greater exposure to green spaces is associated with lower odds of hypertension, obesity, and diabetes. Normalized Difference Vegetation Index (NDVI) in residential areas is negatively correlated with the incidence of metabolic syndrome(Patwary et al., 2024).

However, there is conflicting evidence regarding the impact of green space on cardiovascular disease mortality and risk (GRADE: V). Despite heterogeneous study results and low evidence levels, it appears that residential green space exposure has a robust beneficial effect on metabolic health (GRADE: IV-V), warranting further prospective and mechanistic research.

#### 4. Tumor

Research on green space and cancer primarily focuses on lung cancer, while studies on other cancers (breast, prostate, and skin) suggest green spaces may be protective factors, but overall evidence is very limited due to small cohort sizes(J. Li et al., 2023; Sakhvidi et al., 2022). Additionally, green space may have different impacts on cancer mortality rates for urban and rural residents, with urban residents potentially benefiting more from green spaces(J. Li et al., 2023). The quality of evidence in most current studies is rated as “very low,” indicating the need for higher-quality research to establish the exact relationship between green space and cancer. Given the unclear and highly complex etiology of cancer, along with numerous confounding factors, establishing causation is challenging; thus, future research needs to assess environmental exposure factors and investigate biological mechanisms more precisely.

#### 5. Respiratory and allergic diseases

As previously mentioned, green space exposure may serve as a protective factor against lung cancer. For other chronic non-communicable respiratory diseases, only one study categorized the impact on asthma incidence and COPD incidence and mortality rates for the general population. The results indicated a significant association where an increase of 0.1 in NDVI was linked to reduced asthma incidence, lung cancer incidence, and mortality risk for chronic obstructive pulmonary disease(Mingcheng Tang, Wei Liu, Haifang Li, & Fengyi Li, 2023). Additionally, multiple studies have explored the effects of residential green spaces, including vegetation and parks, on allergic respiratory diseases such as childhood asthma and allergic rhinitis. However, these studies have produced inconsistent results(Cao et al., 2023; Fuertes et al., 2016; K. Lambert et al., 2017; Parmes et al., 2020; Twohig-Bennett & Jones, 2018; X. Wang, Zhou, & Zhi, 2023). Variations in measurement methods of residential green spaces, disease diagnoses, and adjustment for confounding factors across included studies may influence the outcomes. Furthermore, seasonal changes in residential green spaces and their impact on allergens have not been fully considered. Another study involving nine European cohorts suggested an association between residential green spaces and increased childhood asthma and allergic rhinitis. This study emphasized that different types of green spaces, such as coniferous forests, may be associated with increased respiratory disease risks(Parmes et al., 2020).

Collectively, studies examining the impact of green space exposure on respiratory health outcomes indicate inconclusive evidence regarding whether green spaces act as protective factors. The complex interactions between green spaces and respiratory system health may vary across different geographical regions and climatic conditions(Squillacioti et al., 2024). Given the distinct mechanisms underlying chronic non-communicable respiratory diseases, respiratory infections, and allergic diseases, future research should categorically explore these diseases and consider the influence of vegetation types.

The association between residential green spaces and allergic diseases in children and adolescents is an active area of environmental health research. Simultaneously, “child- friendliness” is a focal point in landscape design studies. Certain plant species may act as allergens(Oh, 2022); therefore, in relevant planning and design, careful consideration should be given to the selection of vegetation that could potentially trigger allergic diseases.

#### 6. Pregnancy and neonatal outcomes

According to meta-analysis results, an increase of 0.1 unit in Normalized Difference Vegetation Index (NDVI) is associated with higher birth weight(Ahmer et al., 2024; Akaraci, Feng, Suesse, Jalaludin, & Astell-Burt, 2020; Hu et al., 2021; Lee et al., 2020; Zhan et al., 2020). Additionally, exposure to green spaces is linked to reduced risk of low birth weight (LBW) (Ahmer et al., 2024; Akaraci et al., 2020; Hu et al., 2021; Zhan et al., 2020). While the association between green space exposure and preterm birth (PTB) or small-for-gestational-age (SGA) varies across studies(Akaraci et al., 2020; Hu et al., 2021; Lee et al., 2020; Twohig-Bennett & Jones, 2018; Zhan et al., 2020), these studies also indicate a positive trend in reducing these risks. Some studies suggest a non-linear relationship between green space exposure and birth weight, indicating that moderate levels of green space may be more beneficial than extremely high or low levels(Zhan et al., 2020). The heterogeneity of these research findings suggests the presence of other factors influencing the relationship between green space exposure and pregnancy outcomes, such as socioeconomic variables, other environmental factors, and residential conditions.

#### 7. Mental Health

Residential green spaces are considered a unique and potentially modifiable exposure that can reduce physiological stress and improve mental health(Bratman, Hamilton, & Daily, 2012). The relationship between green spaces and mental health is a multidimensional and complex research area that has garnered increasing attention in recent years. The exact impact of green space exposure on improving mental health outcomes in adults, such as reducing depression(Z. Liu et al., 2023; Roberts, van Lissa, Hagedoorn, Kellar, & Helbich, 2019; Yimin Zhang et al., 2024) and anxiety symptoms, shows high heterogeneity among studies(Z. Liu et al., 2023; Yimin Zhang et al., 2024). Although short-term exposure to natural environments exhibits significant heterogeneity, minor effects suggest a decrease in depressive mood following exposure to natural environments, whereas the increase in green spaces within residential areas alone has limited effects on enhancing positive emotions(W. Yao, Chen, Wang, & Zhang, 2021). However, greener residential environments correspond to higher self-rated health assessments(Twohig-Bennett & Jones, 2018). For specific populations like postpartum depression, the relationship with green spaces is less significant, whereas blue spaces may pose potential risk factors(Cadman et al., 2024). Nevertheless, due to high-risk bias and low-quality studies, the credibility of these results is limited. Future research should aim to reduce biases, enhance study quality, and adhere to reporting guidelines.

Some studies further support the positive impact of green spaces on mental health. Nature-based interventions (NBIs) such as gardening, green exercise, and nature-based therapies have been effective in improving mental health outcomes for adults, including those with existing mental health issues. These interventions include promoting overall mental health through gardening activities(Briggs, Morris, & Rees, 2023; Pan□iru, Ronaldson, Sima, Dregan, & Sima, 2024; Soga, Gaston, & Yamaura, 2017; Spano et al., 2020) and alleviating depression(Briggs et al., 2023). Physical activities in forests have shown improvements in depression(Coventry et al., 2021; Siah et al., 2023; Song et al., 2022), reduction in anxiety(Coventry et al., 2021; Song et al., 2022), enhancement of positive emotions(Coventry et al., 2021; Song et al., 2022), reduction of anxiety symptoms(Coventry et al., 2021; Song et al., 2022), and have been associated with lower cortisol levels(Antonelli, Barbieri, & Donelli, 2019) and systolic blood pressure reduction(Siah et al., 2023). The most effective intervention durations range from 8 to 12 weeks, with optimal dosages varying from 20 to 90 minutes(Coventry et al., 2021). However, it is undeniable that interventional studies may introduce additional placebo effects. This is a significant factor limiting the credibility of the results. A policy review emphasizes the importance of creating psychologically supportive urban environments for adolescents and young adults. It suggests that while cities offer opportunities for medical, educational, and economic benefits, urban environments often pose challenges to mental health. Implementing nature-based solutions within cities through parks and urban green spaces is crucial for enhancing the mental health and well-being of urban residents(Collins et al., 2024).

### 7. Associations between Blue exposure and health outcomes

Blue Spaces refer to all forms of natural and artificial surface water bodies, which are essential components of urban environments. There is currently only one retrieved quantitative analysis of evidence regarding the association between Blue Spaces and health outcomes. The study found that urban Blue Spaces are positively associated with decreased obesity rates, lower all-cause mortality, overall health status, and self-reported psychological health and well-being. Blue Spaces facilitate physical activity and play a significant role in providing restorative environments. The impact of Blue Spaces, including coastlines, on human health in residential environments appears promising, but more evidence is still needed(Georgiou, Morison, Smith, Tieges, & Chastin, 2021).

## Discussion

### Key findings

This review included a total of 46 meta-analyses. Most of these meta-analyses were observational and assessed green space exposure using both objective and subjective parameters, although significant variations existed between studies. Overall, exposure to green spaces showed protective effects on all-cause and cardiovascular disease mortality, overall cardiovascular disease incidence, diabetes and metabolic syndrome, low birth weight, and mental illnesses. Contact with natural environments, including gardening activities, promoted reductions in depression, anxiety, stress, and cortisol levels. Exposure to blue spaces was positively associated with reduced all-cause mortality, overall health status, and self-reported psychological health and well-being.

In contrast, within the included systematic reviews, evidence regarding green space exposure and disease-specific mortality, cancer, asthma, and allergic rhinitis was heterogeneous and remains unclear. AMSTAR2 assessments indicated that most included systematic reviews and meta-analyses had one or more methodological limitations, potentially introducing credibility biases into the synthesized evidence.

### Potential mechanisms underlying greenspace and health

Green spaces and their impact on health outcomes involve potential mediating factors, which we categorize into three aspects for discussion.

Firstly, green spaces provide ecological services themselves(Kumar et al., 2023). They are part of residential environments and influence other environmental factors such as heat exposure, air pollution levels, and noise(X. Yao et al., 2022), which are causally linked to various health outcomes. For instance, 1. High temperatures affect thermoregulation in humans, leading to heat-related illnesses such as heatstroke, heat fatigue, and heat cramps. Long-term exposure to high temperatures increases the risk of cardiovascular diseases(Ebi et al., 2021). 2. Air pollutants like particulate matter (PM2.5 and PM10), nitrogen dioxide, sulfur dioxide, and ozone contribute to respiratory diseases such as asthma, COPD, and lung cancer(Hoek et al., 2013), and are associated with increased incidence and hospitalization rates for cardiovascular diseases(Langrish et al., 2012). 3. Noise can cause increased psychological stress(Stansfeld, Haines, Burr, Berry, & Lercher, 2000; Zaman, Muslim, & Jehangir, 2022) and elevated risk of cardiovascular diseases(Münzel, Sørensen, & Daiber, 2021). Environmental factors impact health in multiple ways, through direct physiological effects and by influencing behaviors and mental health. Therefore, reducing exposure to these environmental risk factors is crucial for protecting public health.

Secondly, green spaces benefit residents’ health by providing social and activity settings (Coombes, Jones, & Hillsdon, 2010). Green (and blue) spaces serve as platforms that promote physical activity and social interaction(Astell-Burt et al., 2022; Gascon, Zijlema, Vert, White, & Nieuwenhuijsen, 2017), encouraging outdoor activities such as walking, exercising, leisure, and socializing. These activities not only promote physical health but also enhance social interactions, improving mental well-being. Exercise improves cardiorespiratory function(Tucker et al., 2022), prevents and manages chronic diseases(Booth, Roberts, & Laye, 2012), enhances bone health(Chang, Xu, & Zhang, 2022), and boosts cognitive function(Sewell et al., 2021); while social interactions provide emotional support, reduce loneliness, and help alleviate stress and anxiety(Collins et al., 2024). Green spaces offer high-quality settings for socializing and activities, which are crucial for their impact on human health.

Additionally, based on our literature review, contact with green (and blue) spaces may have direct effects on residents’ physical health. Activities in green environments can reduce stress and anxiety, improve mood, and enhance psychological well-being(Coventry et al., 2021; Song et al., 2022). Moreover, natural elements in green environments such as trees, water bodies, and vegetation can positively impact physiological indicators like blood pressure(Siah et al., 2023).

The green space indicators covered in this study are as previously described. Most observational studies use objective green space indicators such as NDVI as statistical metrics, which provide convenience in computing statistical measures. Linking residents’ green space exposure to NDVI using satellite imagery has become a research paradigm(De La Iglesia Martinez & Labib, 2023). However, this approach has limitations for many health outcomes. For example, NDVI may overlook information such as vegetation types, health conditions, and biomass within areas, and satellite-derived information is also limited by resolution (Jimenez, Lane, Hutyra, & Fabian, 2022). NDVI values are highly influenced by seasonal changes, and using NDVI values from a single time point may not accurately reflect year-round green space exposure(Holland et al., 2021). This could introduce significant bias in assessing certain diseases such as cardiovascular diseases(Twohig-Bennett & Jones, 2018) and allergic diseases(Cao et al., 2023; Fuertes et al., 2016; K. Lambert et al., 2017; Parmes et al., 2020; Twohig-Bennett & Jones, 2018; X. Wang et al., 2023). Other objective green space indicators, such as proximity to the nearest green space or green space ratio, also suffer from similar information losses. The use of subjective assessments, such as residents’ access to green spaces, may introduce subjective biases. The quality of evidence provided by observational studies is much lower than that of intervention studies, indicating the need for more mixed-method and intervention research in the future.

Future research should pay attention to several important issues. First, there is a risk of information loss when converting and indirectly coupling different green space indicators. Current literature reviews mostly rely on cross-sectional surveys based on NDVI/EVI indices. Therefore, future studies should not be limited to NDVI/EVI but should utilize various indicators reflecting green space. It is essential to enhance the quantitative coupling between green space indicators and ecological functions to deepen our understanding of green space ecological benefits. For example, a recent study using AI-based Google Street View assessed neighborhood features related to coronary heart disease prevalence, highlighting associations between green spaces, forests, and lower CHD incidence(Chen et al., 2024).

Secondly, in causal analysis, it is crucial to distinguish between mediating and confounding factors. Mediating factors are part of the causal chain, acting as intermediate steps between the causal variable (green space) and the outcome variable (health). Confounding factors, on the other hand, are variables correlated with both the cause and the effect, potentially obscuring or masking the true relationships. Factors such as gender, age, education level, regional economic status, and other environmental factors unrelated to green space should be thoroughly analyzed in layers when discussing these confounding factors to clarify causality.

Thirdly, many mediating factors themselves directly influence health independently of the presence of green space. It is important to further clarify the mediating effects reflected by different indicators on various health outcomes and assess their robustness to avoid biases and heterogeneity, thus guiding real-world urban design and landscape planning practices.

Lastly, pure environmental ecological studies can only assess associations and mediating factors but cannot evaluate the physiological reasons behind causal relationships. Future research should integrate knowledge and techniques from multiple fields such as biology, ecology, genetics, molecular biology, and computer science under interdisciplinary backgrounds to delve into the complex mechanisms of human-environment interactions in urban living environments.

### Strengths and limitations

Based on our understanding, this is the second comprehensive review of evidence linking green spaces and human health, summarizing and evaluating the evidence using systematic reviews and meta-analyses. This tertiary-level research surpasses primary and secondary studies in terms of evidence hierarchy. The authors conducted comprehensive searches across three international databases to identify relevant systematic reviews, with study selection and data extraction performed independently by two authors. They strictly adhered to the Preferred Reporting Items for Systematic Reviews and Meta-Analyses (PRISMA) guidelines and assessed the methodological quality of included articles using the AMSTAR2 checklist. This provided an objective assessment of current evidence, identifying gaps and limitations in existing literature and offering recommendations for future systematic reviews. Compared to the initial umbrella review in 2021, we observed progress and standardization in evidence assessment, including improved definitions of health outcomes, enhanced consideration of evidence grading, and expanded evaluation of blue spaces. Particularly, detailed discussions on mediating factors were added to provide guidance for future green space planning and development.

However, there are several issues to note:

AMSTAR2 assessments indicated methodological limitations in most included meta-analyses, potentially undermining the credibility of synthesized evidence.

Umbrella reviews can only synthesize associations between green spaces and health outcomes reported in published systematic reviews, potentially missing or underestimating associations not covered in these reviews.

Most studies are based on large-sample cross-sectional observations, which objectively rely on vegetation indices like NDVI/EVI derived from satellite imagery within various buffer zones. This approach has significant limitations in seasonal-related health outcomes (such as increased cardiovascular disease rates in winter and allergic diseases in spring) and in distinguishing specific vegetation types and species, which are important sources of bias.

There remains considerable heterogeneity in health outcome assessments across studies, and pure environmental ecology alone cannot explain the sources of this heterogeneity.

### Concluding remarks and future perspectives

Overall, we observe that exposure to green spaces has a protective effect on overall mortality and cardiovascular disease mortality, cardiovascular disease incidence, diabetes and metabolic syndrome, low birth weight, and mental disorders. Contact with natural environments, including gardening activities, promotes reduction in depression, anxiety, stress, and cortisol levels. Exposure to blue spaces is positively associated with reduced overall mortality, improved overall health status, and self-reported psychological well-being and happiness. However, the impact on other health outcomes is limited or uncertain. Nevertheless, these findings primarily derive from heterogeneous cross-sectional studies. Therefore, there is a need for longitudinal or intervention study designs to examine causality; there is a requirement for more accurate quantitative assessments targeting mediating factors. For instance, dynamic assessments of green exposure using big data technologies should be applied, taking into account residents’ utilization of green facilities. Studies should include more populations from low- and middle-income countries. Additionally, future systematic reviews should strictly adhere to standard guidelines to enhance their methodological quality.

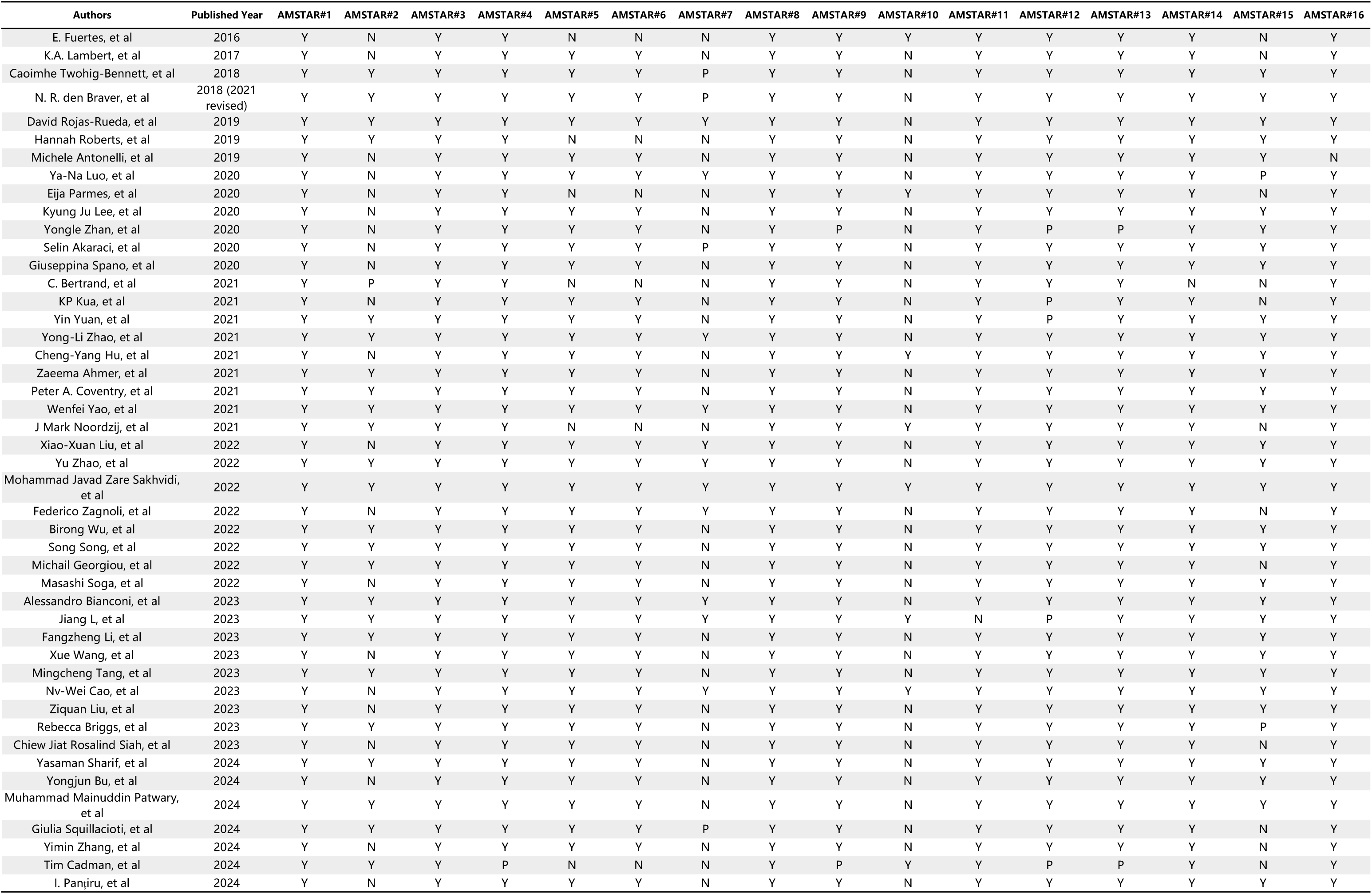
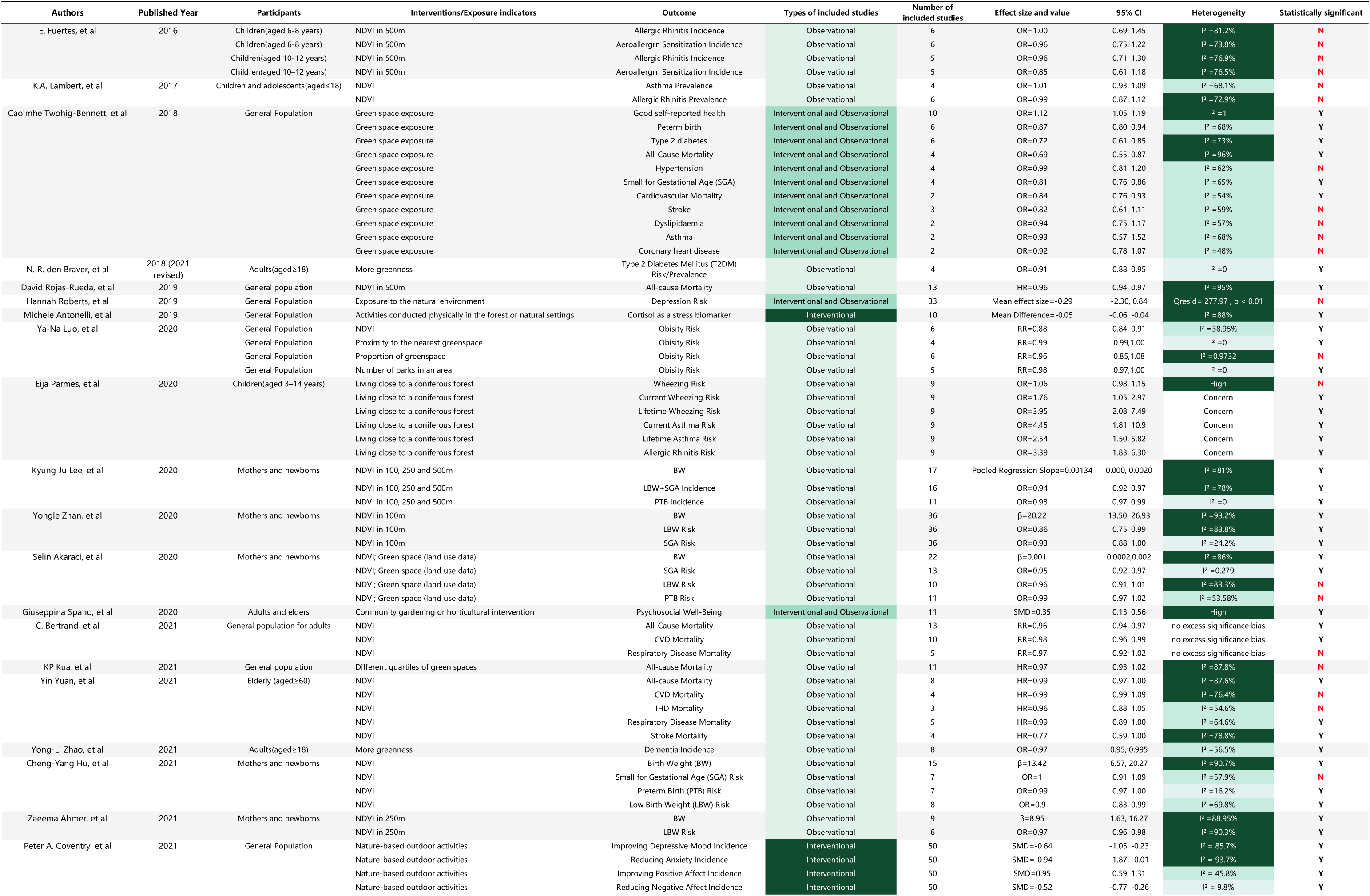
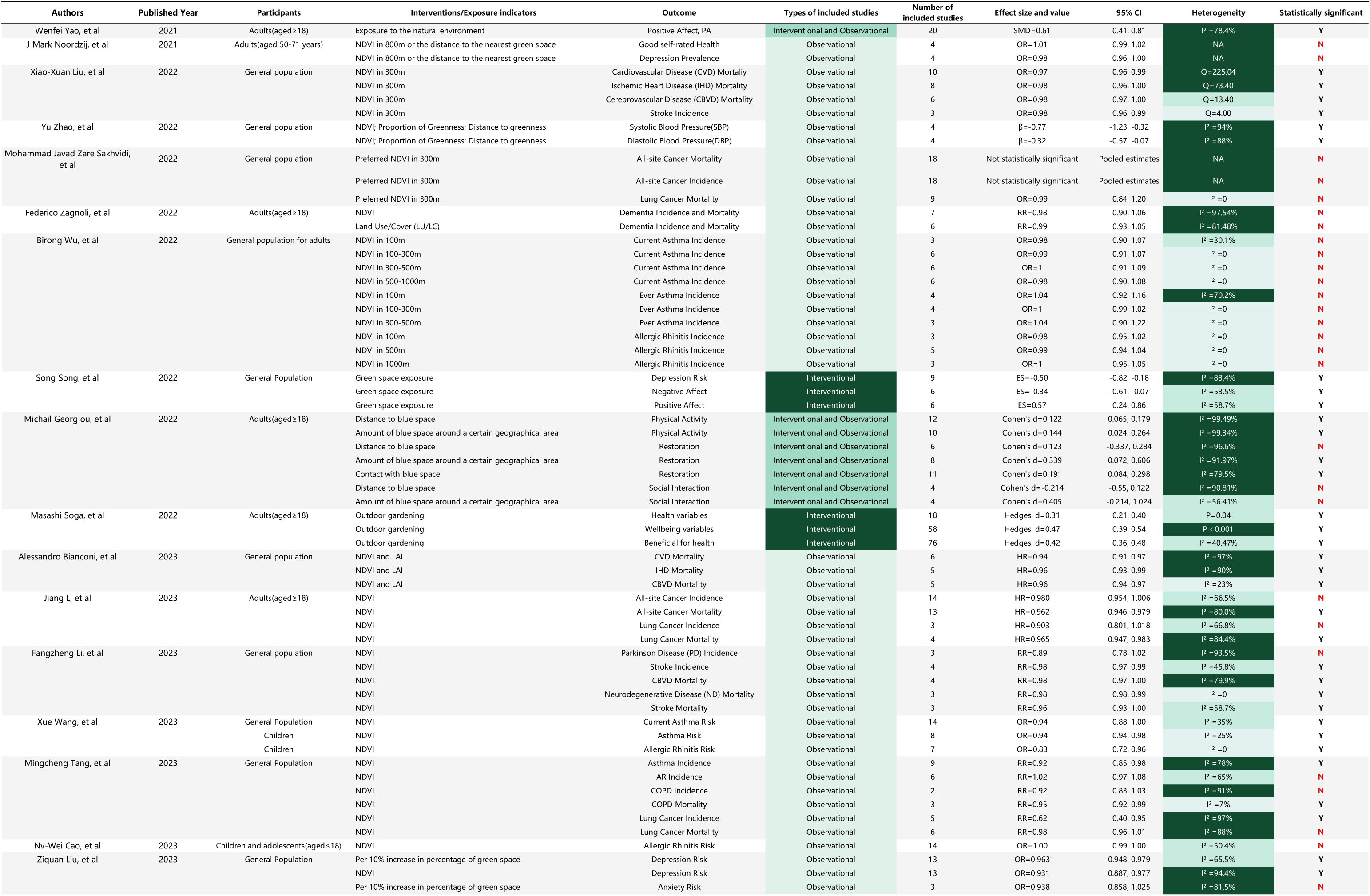
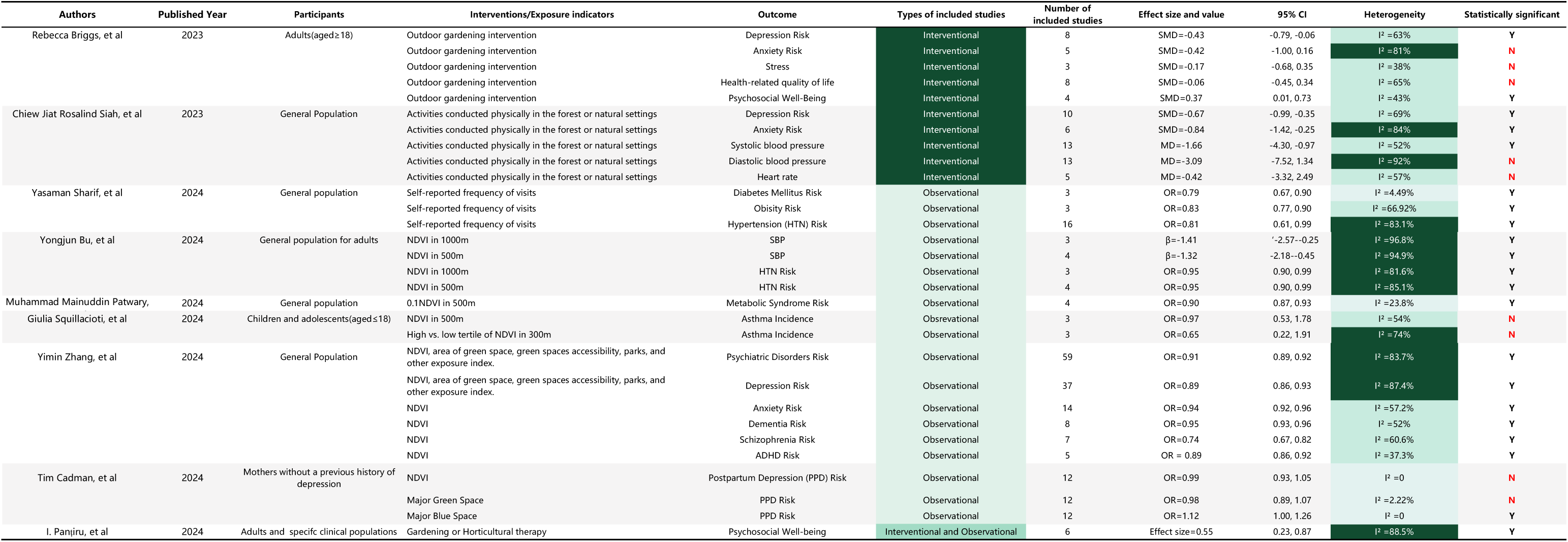
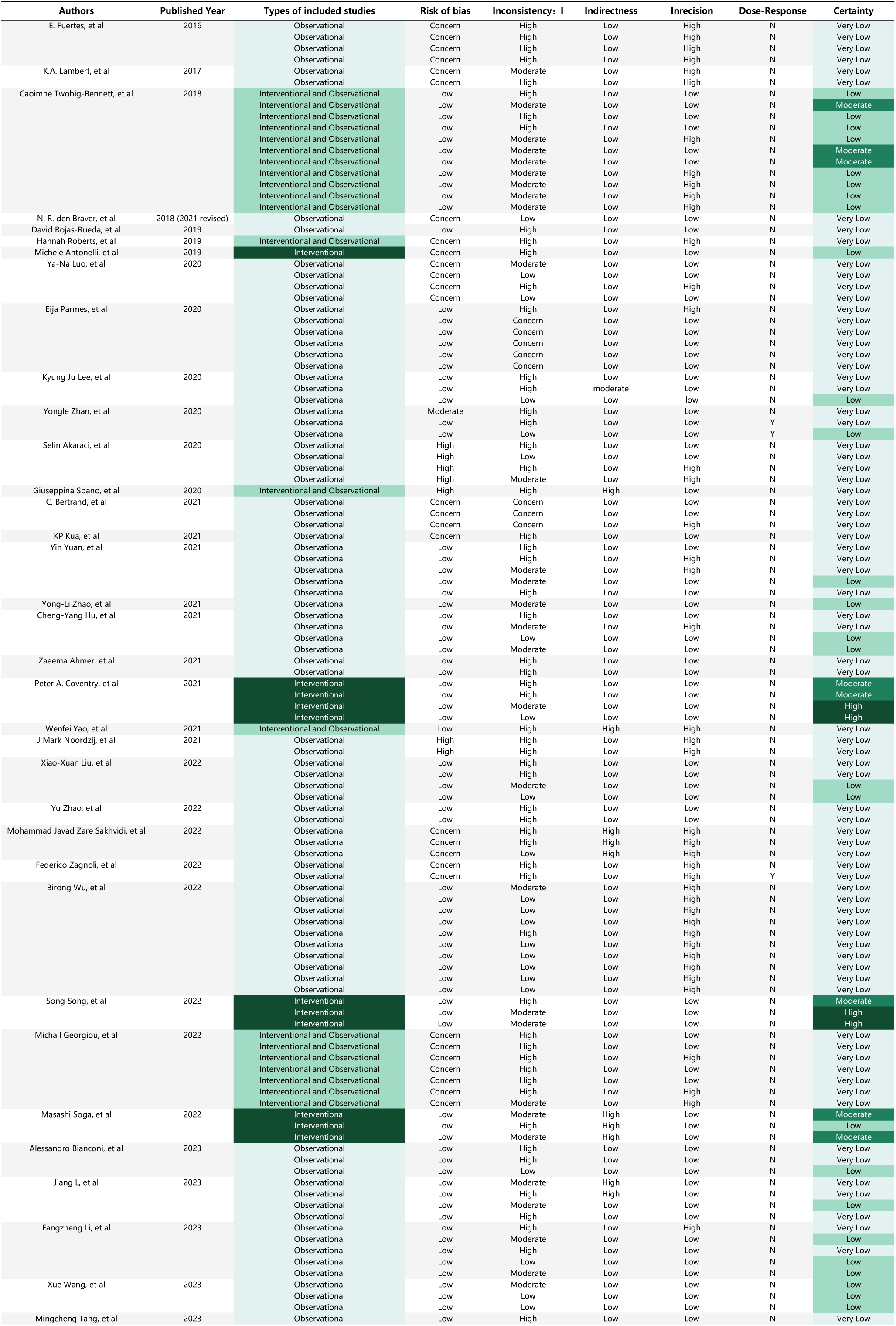
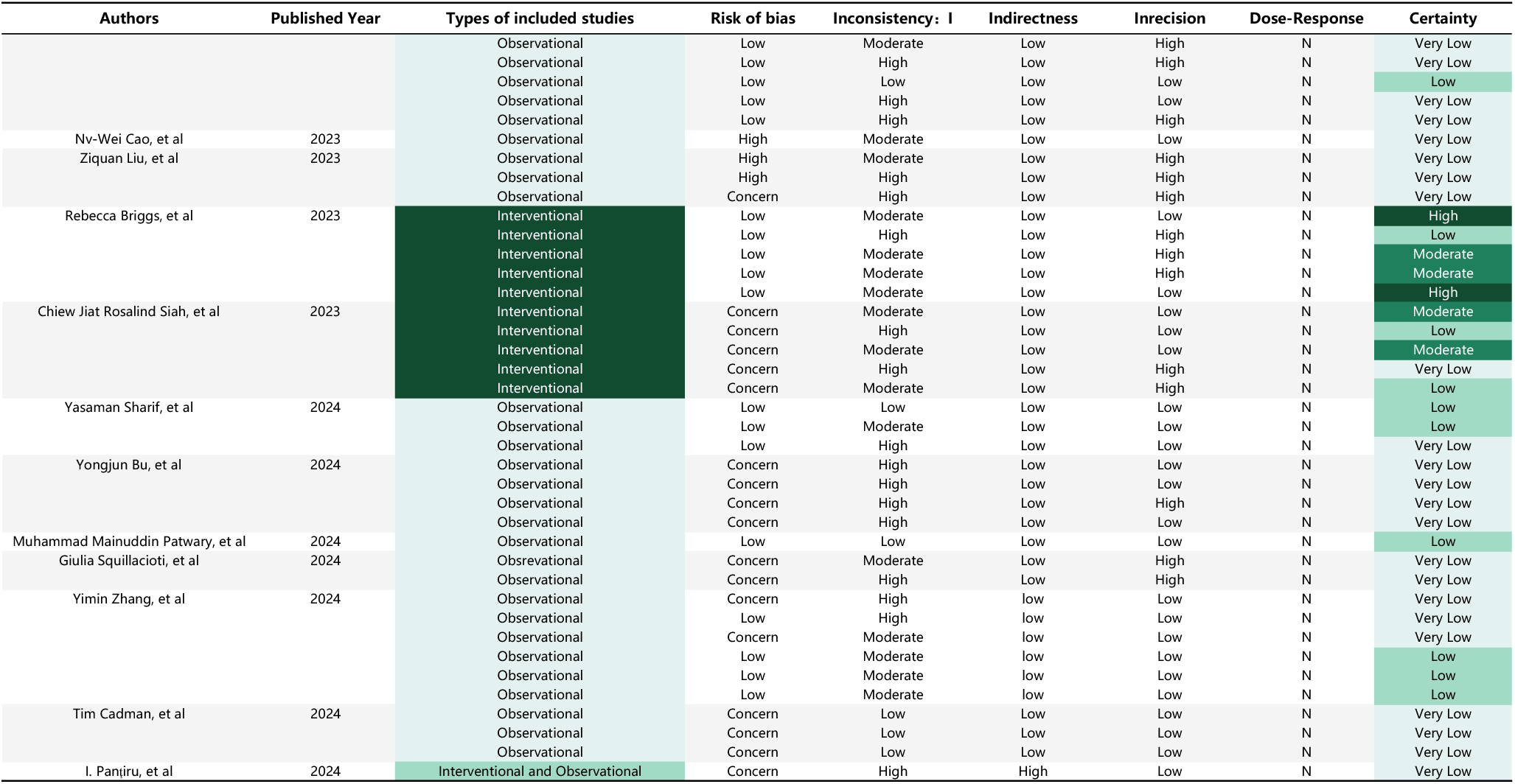

## Data Availability

All data produced in the present work are contained in the manuscript.

